# Wealth inequalities in timely postnatal care in Malawi: how much do antenatal care and place of delivery explain?

**DOI:** 10.64898/2026.11.09.26367675

**Authors:** Redson Mwandama, Spy Munthali, Margubur Rahaman, Joe Maganga Zonda, Rekha Gupta, Salim A Mapila

## Abstract

**Background:** Postnatal care (PNC) is important for maternal and newborn health, but socioeconomic inequalities may limit timely access. In Malawi, evidence on PNC timing and the pathways linking household wealth to PNC is limited. We examined socioeconomic inequalities in timely PNC and whether antenatal care (ANC) and delivery-facility type accounted for part of the wealth gradient.

**Methodology:** We analysed 4,726 women with a live birth in the previous 24 months from the 2024 Malawi Demographic and Health Survey. PNC was timely when the first check occurred within two days of birth, delayed when it occurred later, and absent when no qualifying check was reported. Survey-weighted multinomial logistic regression examined associations between household wealth and PNC timing, with sequential adjustment for maternal education, ANC and place of delivery. The slope index of inequality (SII) and relative index of inequality (RII) summarised inequality. Pathway analyses examined ANC and delivery-facility type as mediators.

**Results:** Overall, 66.6% of women received timely PNC, 5.1% delayed PNC and 28.3% no PNC. Unadjusted, women in the richer and richest wealth quintiles had lower relative risk ratios (RRR) of no PNC than women in the poorest quintile (RRR 0.56 and 0.57, respectively). After full adjustment, the richer group remained less likely to have no PNC (RRR 0.67, 95% CI 0.50–0.89). Wealth inequality persisted (SII −8.4 percentage points, 95% CI −14.8 to −2.0; RII 0.74, 95% CI 0.59–0.93). Among all births, 26.6% of the wealth association with no PNC was accounted for by ANC and place of delivery, primarily home delivery.

**Conclusion:** Timely PNC remains unequally distributed by household wealth in Malawi. The findings suggest that reducing missed PNC opportunities requires attention to socioeconomic barriers and continuity from pregnancy through delivery and the postnatal period.

## Introduction

Postnatal care (PNC), a crucial part of the maternal and newborn health continuum, remains largely overlooked despite the high concentration of preventable maternal and neonatal deaths occurring in the period immediately following childbirth(1–3) . The first six weeks after birth offer a critical window to identify and manage complications, and to provide essential counseling and support on breastfeeding, family planning, and newborn care (4–6). However, health systems typically prioritize antenatal and intrapartum care, allocating fewer resources and less attention to postnatal services. This imbalance is also reflected in the limited research available on PNC (4). Evidence from low- and middle-income countries consistently reveals that contact with maternity services during pregnancy and childbirth often fails to translate into timely postnatal care.

For instance, in Bangladesh, PNC from a medically trained provider within two days increased from 16% in 2004 to 52% in 2017 (7). Yet, significant disparities persisted, with women from the poorest households, those delivering at home, and those who received no antenatal care (ANC) being the least likely to receive PNC (7,8). A pooled analysis of 23 countries similarly found that only 41.4% of women received quality maternal PNC, with higher ANC attendance and facility delivery correlating with increased use (9). Ethiopia reinforces this trend: only 16.5% of women received PNC within two days (10). Women with at least four ANC visits had substantially higher odds of timely PNC (aOR 2.50, 95% CI 1.42–4.42), and institutional delivery was strongly associated with timely care (aOR 14.55, 95% CI 2.21–95.77)(10) . In rural Tanzania, only 14.6% of women sought PNC within 48 hours (11). Qualitative data pointed to limited knowledge, inadequate provider scheduling, workload, and service constraints as additional barriers to postnatal care (11). Collectively, these studies indicate an incomplete transition from ANC and delivery to PNC, highlighting the relevance of both socioeconomic status and prior engagement with maternal health services in determining whether women receive timely care.

However, the precise relationship between these factors is less clear. While evidence consistently links wealth to PNC use (1,12), and ANC and facility delivery to subsequent postnatal contact (6,10), these factors may represent interconnected stages rather than independent determinants. A woman’s economic position can influence her ability to access ANC and deliver at a facility. Concurrently, ANC contact can provide crucial information and establish a relationship with the health system prior to childbirth. Delivering in a health facility then offers an immediate opportunity for a postnatal check before discharge (6,10,13). The existing literature has largely examined these associations in isolation, making it challenging to ascertain how much of the socioeconomic disparity in PNC is explained by earlier care received in the maternity pathway (14). This distinction is particularly important where facility delivery is already prevalent, as a persistent wealth gradient among facility births would suggest that inequalities extend beyond mere physical access to a delivery facility.

The 2024 Malawi Demographic and Health Survey reported that 64.8% of women with a recent live birth received a postnatal check within two days of delivery (15). Coverage increased from 58.9% in the lowest wealth quintile to 68.1% in the highest (15). The difference was even more pronounced by place of delivery: 66.3% of women who delivered in a health facility received a check within two days, compared with 18.5% among those who delivered elsewhere (15). Earlier studies also documented gaps in postnatal care within the country. For instance (16) reported that 48.4% of women received skilled PNC within 42 days in the 2015–16 MDHS, while (17) found that only 3% of rural women received maternal PNC within 24 hours. Facility-based evidence from Thyolo demonstrated a decline in postnatal attendance from 90.5% within 48 hours to 30.2% at three to seven days and 6.1% at eight to 42 days (18). Although these studies employ different measures and timeframes, they consistently point to persistent gaps in postnatal contact, especially among women who do not deliver in health facilities.

Despite this evidence, the socioeconomic pattern in timely PNC in Malawi has not been examined as a sequence of connected stages of care. Previous studies have identified ANC attendance, residence, employment, mode of delivery and delivery-facility type as correlates of PNC use (16,19,20), but they have generally treated PNC as a binary outcome or used a single timing threshold. It therefore remains unclear whether household wealth distinguishes women who receive PNC on time from those who receive it late or not at all, and to what extent differences in ANC attendance and place of delivery account for this pattern. It is also important to establish whether the wealth gradient persists among women who have already delivered in a health facility. This study examines socioeconomic inequalities in the timing of PNC in Malawi, distinguishing timely, delayed and absent care, and assesses the extent to which ANC attendance and delivery-facility type account for the observed wealth differences.

## Methods

### Study design, data sources, and participants

We did a cross-sectional analysis of the 2024 Malawi Demographic and Health Survey (MDHS), a nationally representative household survey. The survey was implemented by the National Statistical Office with technical assistance from ICF through The DHS Program. Fieldwork took place from 13 May to 31 August 2024. The survey used a stratified two-stage cluster design, selecting 792 clusters and about 23,760 households. We used the Women’s Individual Recode file, which holds the birth history and the maternal and newborn care modules. Eligible women were aged 15–49 years and had a live birth in the 24 months before interview. For women with more than one such birth, we analysed the most recent. We excluded records with a zero sampling weight, which removes the Dzaleka refugee-camp domain that the MDHS reports separately. We then excluded women with missing data on any model variable (n=27) and women who reported a check by a qualified provider but whose timing was unknown or missing (n=88). The analytic sample comprised 4,726 women, of whom 4,593 delivered in a health facility (S1 Table). All participants were women by design, so we did not analyse by sex or gender. We used anonymised, publicly available secondary data from The DHS Program, which were accessed for research purposes on 20 September 2026. The authors had no access to information that could identify individual participants during or after data collection. Ethical approval and informed consent for the survey were obtained by the survey implementers, so no further ethical approval was required for this analysis. We followed the STROBE guideline for cross-sectional studies.

### Procedures

The outcome was the timing of the first postnatal check, in three mutually exclusive categories. A check counted if it was done by a doctor, clinical officer or medical assistant, a nurse or midwife, or a patient attendant. Checks by other providers, including traditional birth attendants and community health workers, were treated as no check. We took the earlier of the check before discharge from the facility and the check after discharge or after a home delivery as the first check, and converted its timing to hours since delivery. Timely PNC was a first check during the first two days after birth (reported at under 72 hours, or at two days or less, following the DHS definition). Delayed PNC was a first check later than this, including after 41 days. No PNC was no qualifying check at any time. We separated delayed from no PNC because they reflect different failures of care: contact that comes too late, and contact that never happens.

The main explanatory variable was household wealth, measured by the DHS wealth quintile (poorest to richest). Maternal education was categorised as none, primary, secondary or higher. ANC use was the number of visits for the most recent birth (none, 1–3, or four or more). Delivery-facility type had four categories: public, CHAM or mission, private (NGO and non-NGO), and other and home delivery. In analyses of facility births, we used the first three categories. All models adjusted for maternal age at birth (under 20, 20–34, 35–49 years), birth order (1, 2–3, 4–5, 6 or more), marital status (married or living with a partner; formerly married, which includes widowed, divorced and separated; never married), urban or rural residence, and region (Northern, Central, Southern). Caesarean delivery was described but not used in the models, because it follows the choice of facility and may lie on the pathway.

### Statistical analysis

All analyses accounted for the complex survey design. Each woman *i* carried a weight *w*_*i*_ (the DHS individual weight divided by 10^6^), a primary sampling unit and a stratum, and variances were estimated by Taylor linearisation, with single-PSU strata centred. We first estimated the weighted proportion of women in each PNC category *j* within each subgroup *g*, with 95% CIs:

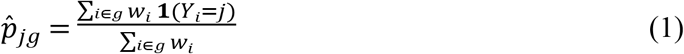

Percentages in the descriptive tables are weighted and counts are unweighted.

Associations between each characteristic and PNC timing were tested with the design-based F test (Rao–Scott corrected Pearson). We did this for all births and for facility births. To check the outcome coding, we compared the weighted proportion with a first check within two days, and with no check, against the published MDHS estimates. The weighted proportion with a first check within two days (65.4%, 95% CI 63.5 to 67.3) and with no check (28.0%, 26.3 to 29.7) matched the published MDHS estimates (64.8% and 27.9%). We then fitted survey-weighted multinomial logistic models for PNC timing *Y*_*i*_ ∈ {1,2,3} (1 timely, 2 delayed, 3 none), with timely PNC as the reference:

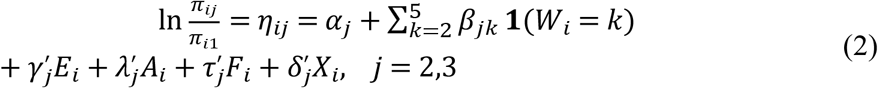

where π_*ij*_ = Pr(*Y*_*i*_ = *j*∣ ⋅) = exp(η_*ij*_)/∑_*m*_ exp(η_*im*_) with *η*_*i*1_ = 0, *W*_*i*_ is the wealth quintile (poorest as base), *E*_*i*_ education, *A*_*i*_ ANC category, *F*_*i*_ delivery-facility type, and *X*_*i*_ the covariates. We estimated the parameters by weighted pseudo-maximum likelihood, maximising ∑_*i*_ *w*_*i*_∑_*j*_ **1**(*Y*_*i*_ = *j*)lnπ_*ij*_. We fitted four nested models. Model 1 included wealth and *X*_*i*_ only. Model 2 added *E*_*i*_, Model 3 added *A*_*i*_, and Model 4 added *F*_*i*_. We report relative risk ratios, RRR_*jk*_ = exp *β*_*jk*_, and tested the joint null *β*_*jk*_ = 0 for all quintiles and both outcomes with a design-based Wald test. Wealth was also entered as a linear term, per one-quintile step, in Models 1 and 4 to test for a trend. The same four models were fitted for facility births, with three facility types. To show the gradient on the probability scale, we computed marginally standardised predicted probabilities for each wealth quintile *k* from Models 1 and 4:

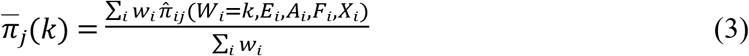

Coefficients from nested non-linear models are rescaled when covariates are added, so changes in *β*_*jk*_ across Models 1 to 4 are not a clean measure of attenuation. We therefore defined two binary outcomes, *Y*^1^ = 1 for no PNC versus any PNC, and *Y*^2^ = 1 for not timely (delayed or none) versus timely. For each, we fitted nested survey-weighted logistic models with the same covariate sequence, Pr(*Y*_*i*_ = 1) = Λ(η_*i*_), where *Λ* is the logistic function. From each model we computed the average marginal effect of wealth quintile *k* compared with the poorest:

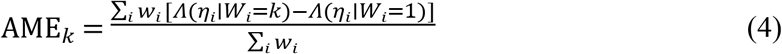

Because AMEs are on the probability scale, they are comparable across models. To obtain confidence intervals for attenuation, we refitted the four nested models as survey-weighted linear probability models and combined them with Stata’s suest command. We then used the delta method to estimate the percentage reduction in each wealth difference between Model 1 and Model 4, 100 × (*b*_1_ − *b*_4_) / *b*_1_, and the share of that reduction due to each added block (education, ANC, delivery place). To summarise the gradient in one number, we ranked women by the DHS wealth index score using the survey weights, giving each woman a fractional rank from 0 (poorest) to 1 (richest). We estimated the slope index of inequality (SII), the absolute difference in probability between the richest and poorest, as the coefficient on this rank in a survey-weighted linear model. We estimated the relative index of inequality (RII), the corresponding ratio, from a survey-weighted Poisson model with a log link. Both were estimated for each binary outcome in each of the four nested models. To estimate direct and indirect pathways, we fitted a survey-weighted generalised structural equation model (GSEM) with three linked equations, corresponding to the pathway wealth → ANC → delivery-facility type → PNC timing. Wealth *W*_*i*_ and ANC *A*_*i*_ entered the downstream equations as linear scores, so wealth effects are per one-quintile step. ANC was modelled by ordered logistic regression:

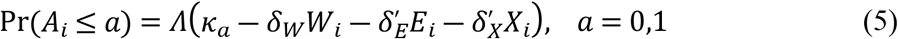

Delivery-facility type was modelled by multinomial logistic regression with public facilities as the base:

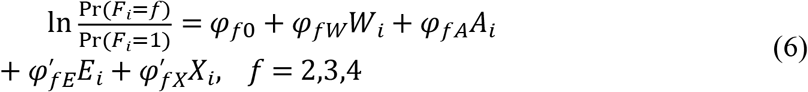

PNC timing was modelled by multinomial logistic regression with timely PNC as the base:

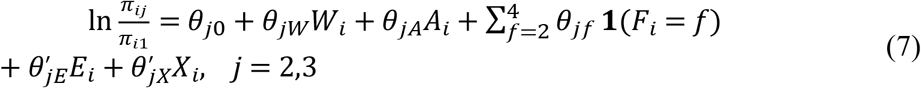

From these we derived, on the log relative-risk scale and for each outcome *j* versus timely PNC, the following effects of wealth:

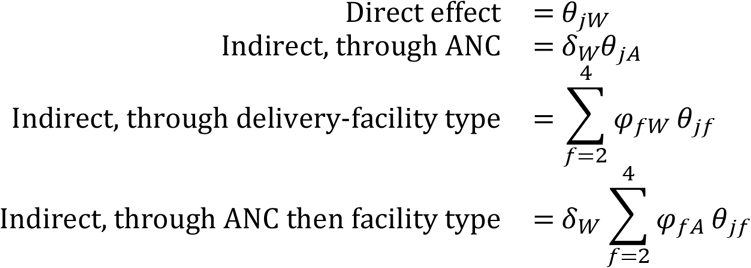

The total indirect effect is the sum of the three indirect paths, and the total effect is the direct effect plus the total indirect effect. We obtained 95% CIs by the delta method, Var 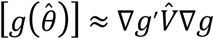, where 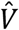 is the design-based covariance matrix of the estimates. Products of coefficients from non-linear equations approximate mediated effects on the log scale, so we interpret them as indicative of pathways and not as exact decompositions. Delayed PNC accounted for about 5% of births, and the “other and home” category was small (133 women) and almost fully determined no PNC. We therefore also estimated the pathways on the probability scale with linear path models, for each binary outcome *Y* ∈ {*Y*^1^,*Y*^2^}:

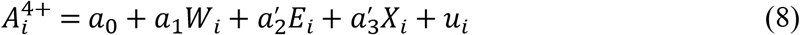

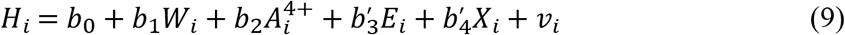

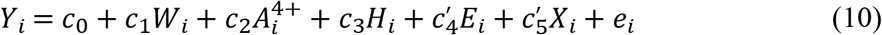

where 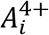 indicates four or more ANC visits and *H*_*i*_ indicates other and home delivery (all births) or non-public facility delivery (facility births). With linear equations, the direct effect is *c*_1_. The indirect effects are *a*_1_*c*_2_ (through ANC), *b*_1_*c*_3_ (through delivery place) and *a*_1_*b*_2_*c*_3_ (through ANC then delivery place). The total effect is *c*_1_ plus the three indirect effects, and the proportion mediated is the total indirect effect divided by the total effect. We report the proportion mediated only where the total effect was clearly different from zero.

Confidence intervals again used the delta method. In sensitivity analyses, we repeated the analysis with timely PNC defined as a first check before 48 hours (responses of two days counted as not timely), and with a first check after 41 days classed as no PNC, following the DHS rule. Both used the unadjusted wealth model and the fully adjusted model. All tests were two-sided with 95% CIs. We did all analyses in Stata version 19.5.

### Conceptual framework

We analysed the timing of postnatal care through a conceptual framework linking household wealth to PNC through the maternity services that precede it (Fig 1). The framework draws on the WHO social determinants of health framework, the continuum-of-care concept, Andersen’s behavioural model of health service use, and the three-delays model (21–24). Together, these perspectives suggest that socioeconomic position shapes access to maternal health services and that contact with ANC and delivery services may influence subsequent PNC. Household wealth is the main exposure and may influence timely PNC through ANC utilization, delivery-facility type and direct socioeconomic constraints.

**Fig 1.**
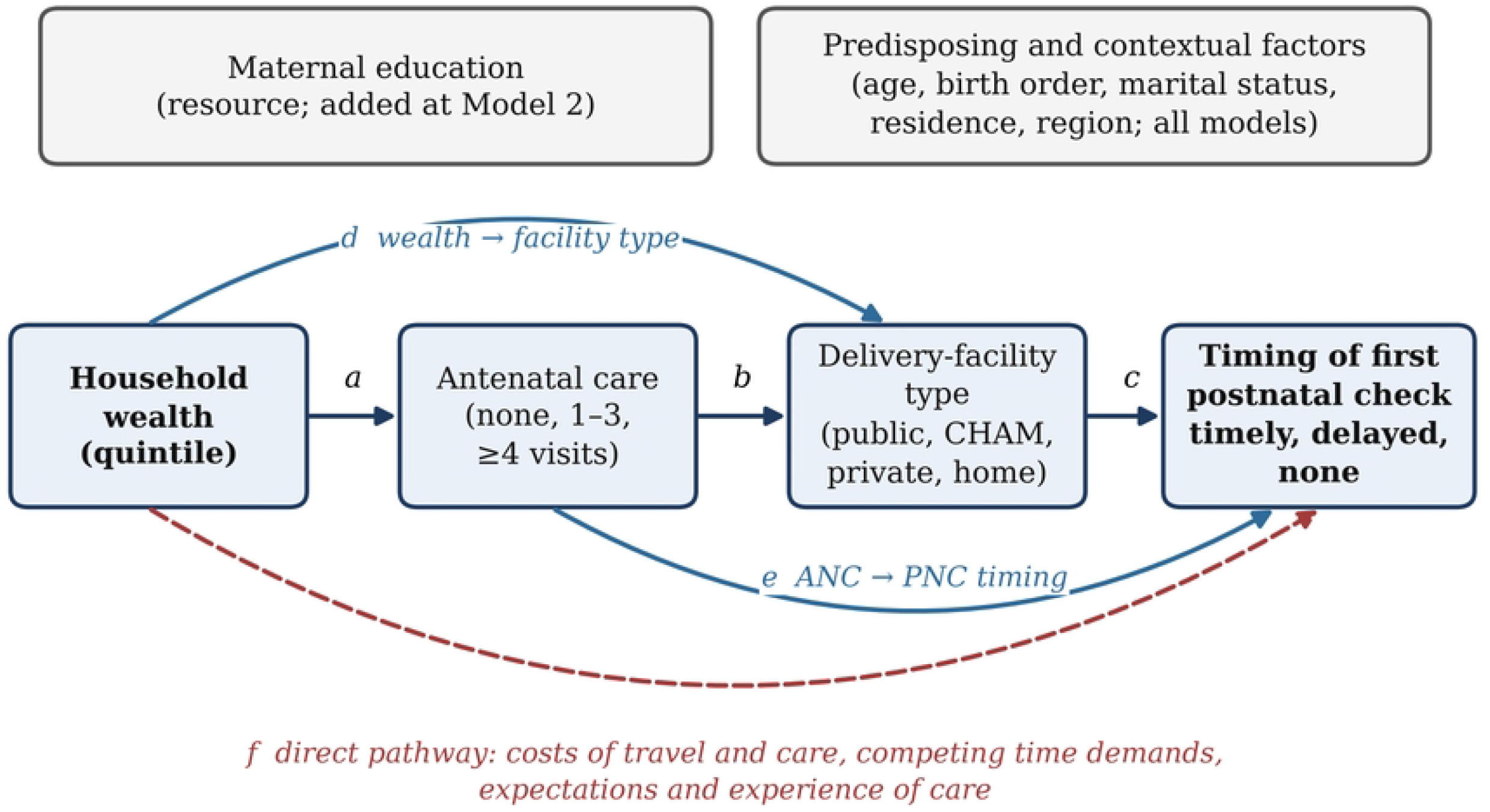
Conceptual framework linking household wealth to the timing of postnatal care. Paths a to c show the sequential pathway through ANC and delivery-facility type, paths d and e skip one stage, and path f is the direct pathway not explained by ANC or facility type. These paths correspond to the structural equation model in equations 5 to 7. Maternal education enters the nested models at Model 2, and the other covariates enter all models. ANC=antenatal care. CHAM=Christian Health Association of Malawi. PNC=postnatal care.

Wealthier women may have greater ability to attend ANC, access different types of delivery facilities and meet the costs associated with seeking care. ANC may provide information about postnatal care and establish contact with health services before delivery, while delivery-facility type may influence the opportunity to receive a postnatal check before discharge. Wealth may also remain associated with PNC timing after accounting for these factors through transport costs, competing demands and other barriers to care. Maternal education is considered an additional resource that may account for part of the wealth gradient, while age, birth order, marital status, residence and region are treated as demographic and contextual characteristics.

The framework distinguishes timely PNC, delayed PNC and no PNC because these outcomes represent different points of failure in the care pathway. Delayed PNC indicates that contact occurred but not within the recommended period, whereas no PNC indicates that postnatal care was not initiated. The analysis therefore examines whether the wealth gradient differs across these outcomes and whether it is attenuated after accounting for ANC utilization and delivery-facility type. Among women who delivered in a health facility, the analysis further assesses whether socioeconomic inequalities in timely PNC persist. Because the data are cross-sectional and wealth is measured at the time of interview, the framework represents a conceptual ordering rather than a causal model. The pathway follows the temporal sequence of ANC, delivery and PNC and provides the basis for examining whether differences in ANC utilization and delivery-facility type are associated with the observed wealth gradient in PNC timing.

## Results

### Background characteristics

The analysis included 4,726 women with a recent live birth, of whom 4,593 had delivered in a health facility (Table 1). The sample was predominantly rural (85.9%), and most women were aged 20–34 years (62.6%) and married or living with a partner (77.7%). Primary education was the most common level of maternal education (68.2%), while 22.9% had secondary education and 2.1% had higher education. More than three-fifths of women had attended four or more antenatal care visits (62.6%), while 35.0% had one to three visits and 2.4% had none. Most births occurred in public facilities (83.1%), followed by CHAM/mission facilities (11.3%) and private facilities (2.4%); 3.1% occurred at home or other locations. Caesarean delivery accounted for 11.0% of all births. The distribution of characteristics was similar when restricted to facility births, with only minor differences in the proportions across wealth, education, age, residence, antenatal care and other characteristics.

**Table 1.** Characteristics of the study population, all births and facility births, 2024 Malawi DHS.

| **Characteristic** | **All births** | **Facility births** |
|---|---|---|
|  | *n = 4,726* | *n = 4,593* |
| **Household wealth quintile** |  |  |
| Poorest | 1,090 (26.0) | 1,040 (25.4) |
| Poorer | 825 (19.9) | 803 (19.9) |
| Middle | 863 (18.6) | 839 (18.7) |
| Richer | 918 (17.7) | 899 (17.9) |
| Richest | 1,030 (17.8) | 1,012 (18.0) |
| **Maternal education** |  |  |
| No education | 271 (6.9) | 244 (6.3) |
| Primary | 3,112 (68.2) | 3,025 (68.2) |
| Secondary | 1,219 (22.9) | 1,202 (23.4) |
| Higher | 124 (2.1) | 122 (2.1) |
| **Maternal age at birth (years)** |  |  |
| <20 | 1,153 (25.1) | 1,132 (25.5) |
| 20-34 | 2,983 (62.6) | 2,904 (62.7) |
| 35-49 | 590 (12.3) | 557 (11.9) |
| **Birth order** |  |  |
| 1 | 1,607 (34.0) | 1,584 (34.6) |
| 2-3 | 1,922 (40.5) | 1,881 (40.8) |
| 4-5 | 801 (16.5) | 762 (16.2) |
| 6+ | 396 (9.0) | 366 (8.5) |
| **Marital status** |  |  |
| Married or living with partner | 3,679 (77.7) | 3,569 (77.6) |
| Formerly married | 693 (15.7) | 674 (15.7) |
| Never married | 354 (6.6) | 350 (6.7) |
| **Place of residence** |  |  |
| Urban | 911 (14.1) | 895 (14.3) |
| Rural | 3,815 (85.9) | 3,698 (85.7) |
| **Region** |  |  |
| Northern | 1,032 (13.3) | 1,005 (13.3) |
| Central | 1,523 (41.7) | 1,481 (41.8) |
| Southern | 2,171 (44.9) | 2,107 (44.9) |
| **Antenatal care visits** |  |  |
| 0 visits | 97 (2.4) | 77 (1.9) |
| 1-3 visits | 1,614 (35.0) | 1,554 (34.6) |
| 4+ visits | 3,015 (62.6) | 2,962 (63.4) |
| **Place of delivery** |  |  |
| Public | 3,912 (83.1) | 3,912 (85.8) |
| CHAM/Mission | 585 (11.3) | 585 (11.7) |
| Private | 96 (2.4) | 96 (2.5) |
| Other & home | 133 (3.1) | — |
| **Caesarean section** |  |  |
| No | 4,182 (89.0) | 4,049 (88.6) |
| Yes | 544 (11.0) | 544 (11.4) |

*Note.* Values are unweighted n (survey-weighted column %). Facility births exclude 133 births that took place at home or in other locations

### Timing of first postnatal check

Overall, 66.6% of women received their first postnatal check within two days of delivery, 5.1% received a delayed check, and 28.3% had no recorded postnatal check (Table 2). The distribution of delayed PNC was relatively small across most groups, while the main differences were between timely PNC and no PNC. Timing of PNC differed by household wealth (p=0.002). Timely PNC increased from 60.9% among women in the poorest wealth quintile to 73.0% among those in the richer quintile, while the proportion with no PNC declined from 33.4% to 22.2%. The richest group had 70.3% timely PNC and 25.0% no PNC. Maternal education showed a similar gradient (p<0.001), with timely PNC increasing from 55.6% among women with no education to 73.3% among those with secondary education and 77.6% among those with higher education. No PNC declined from 37.7% to 12.0% across the same groups.

**Table 2.**
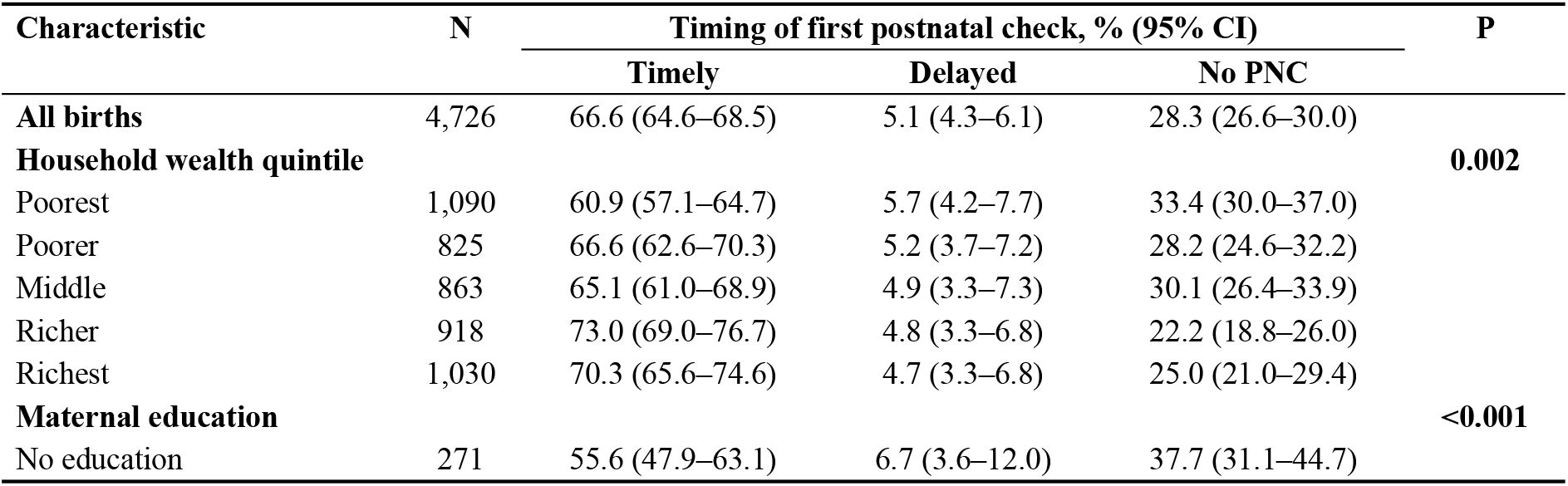
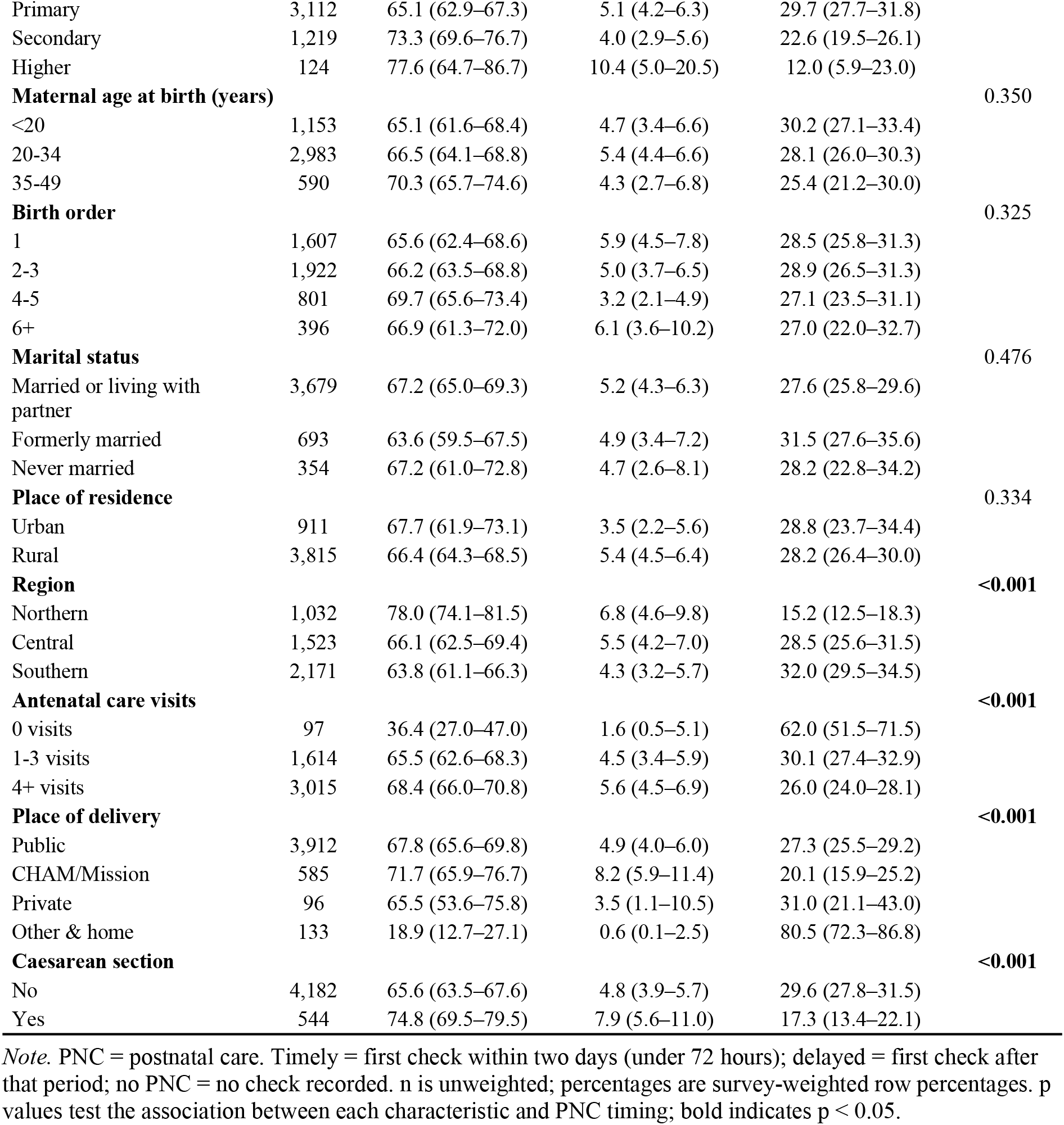
Timing of first postnatal check by background characteristic, all births.

| **Characteristic** | **N** | **Timing of first postnatal check, % (95% CI)** |  |  | **P** |
|---|---|---|---|---|---|
|  |  | **Timely** | **Delayed** | **No PNC** |  |
| **All births** | 4,726 | 66.6 (64.6–68.5) | 5.1 (4.3–6.1) | 28.3 (26.6–30.0) |  |
| **Household wealth quintile** |  |  |  |  | **0.002** |
| Poorest | 1,090 | 60.9 (57.1–64.7) | 5.7 (4.2–7.7) | 33.4 (30.0–37.0) |  |
| Poorer | 825 | 66.6 (62.6–70.3) | 5.2 (3.7–7.2) | 28.2 (24.6–32.2) |  |
| Middle | 863 | 65.1 (61.0–68.9) | 4.9 (3.3–7.3) | 30.1 (26.4–33.9) |  |
| Richer | 918 | 73.0 (69.0–76.7) | 4.8 (3.3–6.8) | 22.2 (18.8–26.0) |  |
| Richest | 1,030 | 70.3 (65.6–74.6) | 4.7 (3.3–6.8) | 25.0 (21.0–29.4) |  |
| **Maternal education** |  |  |  |  | **<0.001** |
| No education | 271 | 55.6 (47.9–63.1) | 6.7 (3.6–12.0) | 37.7 (31.1–44.7) |  |
| Primary | 3,112 | 65.1 (62.9–67.3) | 5.1 (4.2–6.3) | 29.7 (27.7–31.8) |  |
| Secondary | 1,219 | 73.3 (69.6–76.7) | 4.0 (2.9–5.6) | 22.6 (19.5–26.1) |  |
| Higher | 124 | 77.6 (64.7–86.7) | 10.4 (5.0–20.5) | 12.0 (5.9–23.0) |  |
| **Maternal age at birth (years)** |  |  |  |  | 0.350 |
| <20 | 1,153 | 65.1 (61.6–68.4) | 4.7 (3.4–6.6) | 30.2 (27.1–33.4) |  |
| 20-34 | 2,983 | 66.5 (64.1–68.8) | 5.4 (4.4–6.6) | 28.1 (26.0–30.3) |  |
| 35-49 | 590 | 70.3 (65.7–74.6) | 4.3 (2.7–6.8) | 25.4 (21.2–30.0) |  |
| **Birth order** |  |  |  |  | 0.325 |
| 1 | 1,607 | 65.6 (62.4–68.6) | 5.9 (4.5–7.8) | 28.5 (25.8–31.3) |  |
| 2-3 | 1,922 | 66.2 (63.5–68.8) | 5.0 (3.7–6.5) | 28.9 (26.5–31.3) |  |
| 4-5 | 801 | 69.7 (65.6–73.4) | 3.2 (2.1–4.9) | 27.1 (23.5–31.1) |  |
| 6+ | 396 | 66.9 (61.3–72.0) | 6.1 (3.6–10.2) | 27.0 (22.0–32.7) |  |
| **Marital status** |  |  |  |  | 0.476 |
| Married or living with partner | 3,679 | 67.2 (65.0–69.3) | 5.2 (4.3–6.3) | 27.6 (25.8–29.6) |  |
| Formerly married | 693 | 63.6 (59.5–67.5) | 4.9 (3.4–7.2) | 31.5 (27.6–35.6) |  |
| Never married | 354 | 67.2 (61.0–72.8) | 4.7 (2.6–8.1) | 28.2 (22.8–34.2) |  |
| **Place of residence** |  |  |  |  | 0.334 |
| Urban | 911 | 67.7 (61.9–73.1) | 3.5 (2.2–5.6) | 28.8 (23.7–34.4) |  |
| Rural | 3,815 | 66.4 (64.3–68.5) | 5.4 (4.5–6.4) | 28.2 (26.4–30.0) |  |
| **Region** |  |  |  |  | **<0.001** |
| Northern | 1,032 | 78.0 (74.1–81.5) | 6.8 (4.6–9.8) | 15.2 (12.5–18.3) |  |
| Central | 1,523 | 66.1 (62.5–69.4) | 5.5 (4.2–7.0) | 28.5 (25.6–31.5) |  |
| Southern | 2,171 | 63.8 (61.1–66.3) | 4.3 (3.2–5.7) | 32.0 (29.5–34.5) |  |
| **Antenatal care visits** |  |  |  |  | **<0.001** |
| 0 visits | 97 | 36.4 (27.0–47.0) | 1.6 (0.5–5.1) | 62.0 (51.5–71.5) |  |
| 1-3 visits | 1,614 | 65.5 (62.6–68.3) | 4.5 (3.4–5.9) | 30.1 (27.4–32.9) |  |
| 4+ visits | 3,015 | 68.4 (66.0–70.8) | 5.6 (4.5–6.9) | 26.0 (24.0–28.1) |  |
| **Place of delivery** |  |  |  |  | **<0.001** |
| Public | 3,912 | 67.8 (65.6–69.8) | 4.9 (4.0–6.0) | 27.3 (25.5–29.2) |  |
| CHAM/Mission | 585 | 71.7 (65.9–76.7) | 8.2 (5.9–11.4) | 20.1 (15.9–25.2) |  |
| Private | 96 | 65.5 (53.6–75.8) | 3.5 (1.1–10.5) | 31.0 (21.1–43.0) |  |
| Other & home | 133 | 18.9 (12.7–27.1) | 0.6 (0.1–2.5) | 80.5 (72.3–86.8) |  |
| **Caesarean section** |  |  |  |  | **<0.001** |
| No | 4,182 | 65.6 (63.5–67.6) | 4.8 (3.9–5.7) | 29.6 (27.8–31.5) |  |
| Yes | 544 | 74.8 (69.5–79.5) | 7.9 (5.6–11.0) | 17.3 (13.4–22.1) |  |

*Note.* PNC = postnatal care. Timely = first check within two days (under 72 hours); delayed = first check after that period; no PNC = no check recorded. n is unweighted; percentages are survey-weighted row percentages. p values test the association between each characteristic and PNC timing; bold indicates p < 0.05.

Larger differences were observed for antenatal care and place of delivery. Among women with no ANC visits, only 36.4% received timely PNC and 62.0% had no PNC, compared with 68.4% and 26.0%, respectively, among women with four or more ANC visits (p<0.001). Place of delivery was also strongly associated with PNC timing (p<0.001). Timely PNC was reported for 67.8% of births in public facilities and 71.7% of births in CHAM/mission facilities, compared with only 18.9% of births at home or other locations.

Conversely, no PNC was reported for 27.3% of public-facility births and 20.1% of CHAM/mission births, compared with 80.5% of home or other births. Regional differences were also evident (p<0.001). Timely PNC was highest in the Northern Region (78.0%) and lowest in the Southern Region (63.8%), while no PNC was 15.2% in the North compared with 32.0% in the South. In contrast, PNC timing did not differ significantly by maternal age (p=0.350), birth order (p=0.325), marital status (p=0.476), or residence (p=0.334). Caesarean delivery was associated with a higher proportion of timely PNC than vaginal delivery (74.8% versus 65.6%, p<0.001), although this characteristic was not included as an explanatory variable in the subsequent pathway models

### Household wealth and timing of postnatal care

The multinomial models showed that household wealth was primarily associated with whether women received any PNC rather than with delayed versus timely PNC (Table 3).

**Table 3.**
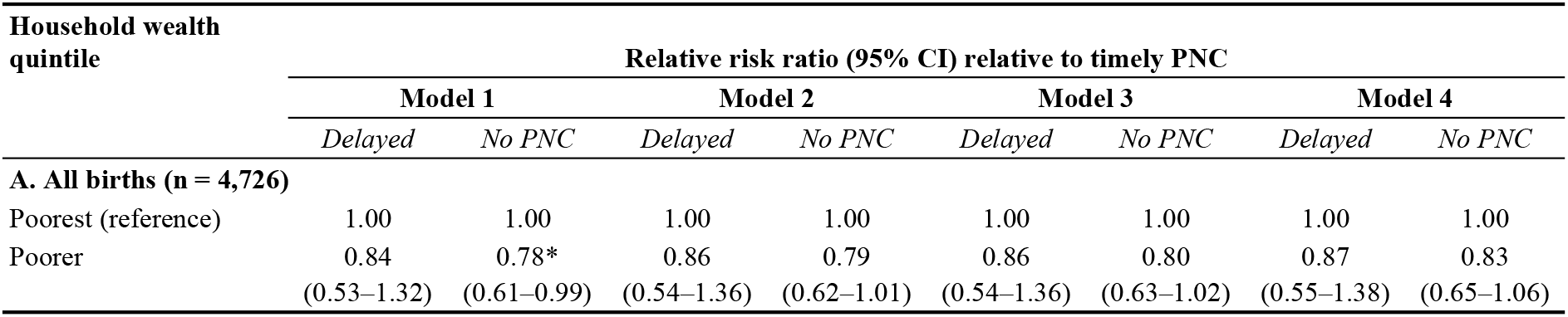
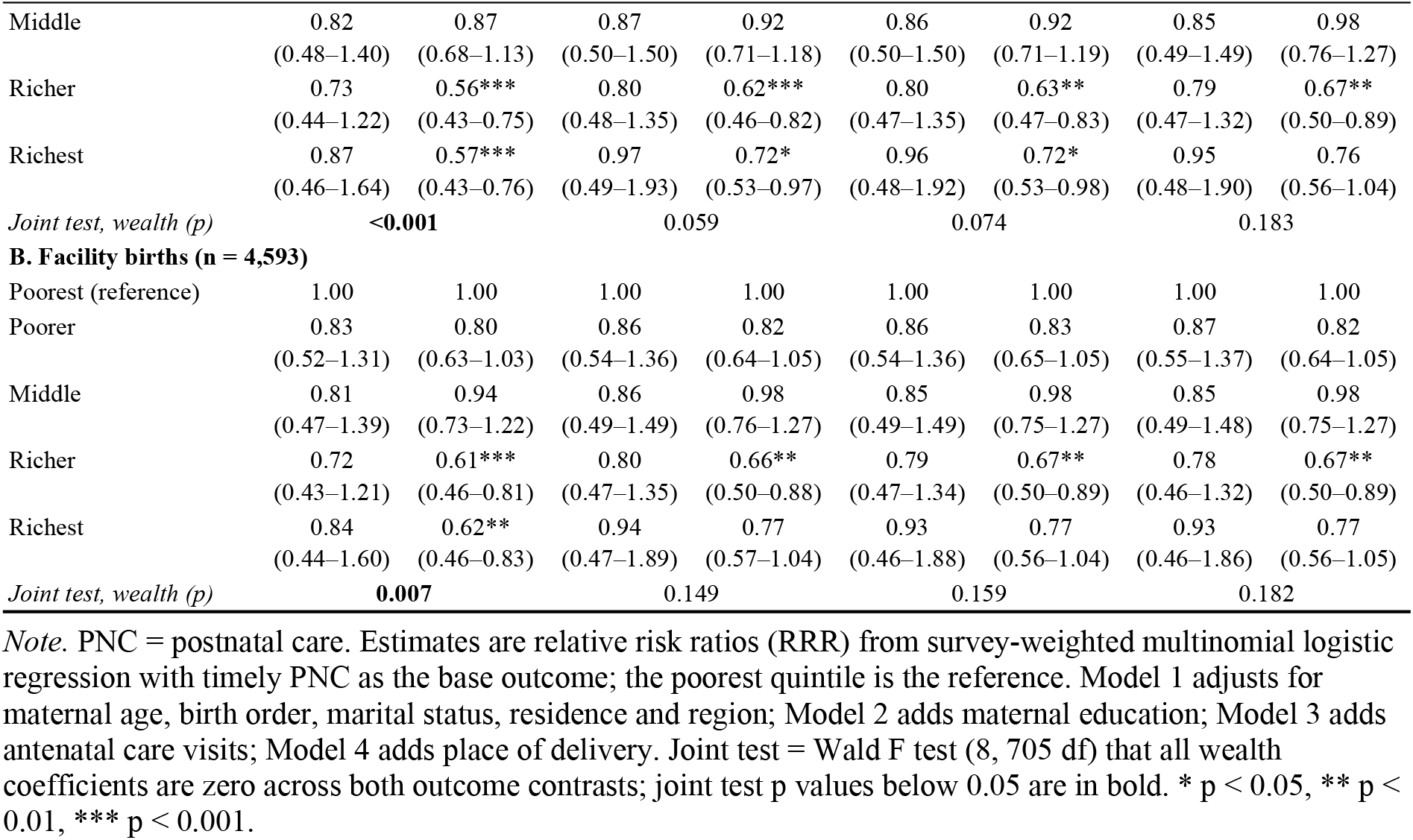
Association between household wealth quintile and timing of postnatal care: relative risk ratios from sequential multinomial models.

Among all births, women in the richer quintile had lower relative risk of no PNC than women in the poorest quintile in Model 1 (RRR 0.56, 95% CI 0.43–0.75), as did women in the richest quintile (RRR 0.57, 95% CI 0.43–0.76). The association was weaker for the poorer quintile (RRR 0.78, 95% CI 0.61–0.99), while none of the wealth groups differed significantly from the poorest group in the delayed PNC contrast. The joint test confirmed an overall association between wealth and PNC timing in Model 1 (p<0.001), driven mainly by differences in no PNC.

Adding maternal education reduced the strength of the wealth associations. The RRR for no PNC was 0.62 (95% CI 0.46–0.82) for the richer group and 0.72 (95% CI 0.53–0.97) for the richest group, while the joint test was no longer statistically significant (p=0.059).

Further adjustment for ANC produced similar estimates, with the richer group remaining less likely to have no PNC than the poorest group (RRR 0.63, 95% CI 0.47–0.83; p<0.01) and the richest group also showing a lower RRR (0.72, 95% CI 0.53–0.98; p<0.05). However, the joint association remained non-significant (p=0.074). After place of delivery was added in Model 4, the association was further attenuated. The richer group remained less likely than the poorest group to have no PNC (RRR 0.67, 95% CI 0.50–0.89), whereas the association for the richest group was no longer statistically significant (RRR 0.76, 95% CI 0.56–1.04).

The overall wealth association was also no longer statistically significant (p=0.183). Across all four models, none of the wealth categories had a statistically significant association with delayed PNC relative to timely PNC.

The facility-birth analysis showed a similar pattern but with weaker overall associations. In Model 1, women in the richer and richest quintiles had lower relative risks of no PNC than women in the poorest quintile (RRR 0.61, 95% CI 0.46–0.81 and RRR 0.62, 95% CI 0.46–0.83, respectively), and the joint wealth test was significant (p=0.007). After adjustment for education, ANC and place of delivery, the association weakened and the joint test was no longer significant (p=0.149, 0.159 and 0.182, respectively). The richer group continued to have a lower relative risk of no PNC in the fully adjusted model (RRR 0.67, 95% CI 0.50–0.89), but the other wealth contrasts were not statistically significant. As in the analysis of all births, wealth was not significantly associated with delayed PNC in any model. The adjusted results indicate that the socioeconomic difference was concentrated in the likelihood of receiving no PNC rather than in delayed receipt among women who received a check. The reduction in the joint wealth association after adding education, ANC and delivery characteristics suggests that these factors account for part of the observed wealth difference, with place of delivery contributing to further attenuation. The persistence of the richer-versus-poorest association in the fully adjusted model, however, indicates that the difference was not completely accounted for by these characteristics.

### Predicted probabilities of postnatal care by household wealth

The predicted probabilities illustrate the magnitude of the wealth differences in PNC timing (Table 4). In Model 1, the predicted probability of timely PNC was 60.9% among women in the poorest quintile compared with 72.5% among those in the richer quintile and 71.7% among those in the richest quintile. Relative to the poorest group, the probability of timely PNC was 11.6 percentage points higher in the richer group (95% CI 6.0–17.2) and 10.8 percentage points higher in the richest group (95% CI 4.6–17.0). In contrast, the predicted probability of delayed PNC varied little across wealth groups, with none of the differences from the poorest group statistically significant. The corresponding differences were more pronounced for no PNC. In Model 1, the predicted probability of no PNC was 33.7% among the poorest women, compared with 22.8% among both the richer and richest groups. This represented differences of −10.8 percentage points (95% CI −16.1 to −5.6) and −10.9 percentage points (95% CI −16.1 to −5.6), respectively. The poorer group also had a lower predicted probability of no PNC than the poorest group, although the difference was smaller (−5.1 percentage points, 95% CI −10.1 to −0.1).

**Table 4.** Predicted probabilities of timely, delayed and no postnatal care by household wealth quintile, Models 1 and 4.

| **Household wealth quintile** | **Model 1** |  |  | **Model 4** |  |  |
|---|---|---|---|---|---|---|
|  | *Timely* | *Delayed* | *No PNC* | *Timely* | *Delayed* | *No PNC* |
| **A. Predicted probability, % (95% CI)** |  |  |  |  |  |  |
| Poorest | 60.9 (57.0–64.7) | 5.5 (3.8–7.2) | 33.7 (30.1–37.2) | 63.5 (59.8–67.2) | 5.4 (3.7–7.1) | 31.1 (27.7–34.5) |
| Poorer | 66.4 (62.5–70.3) | 5.0 (3.3–6.7) | 28.6 (24.8–32.4) | 67.2 (63.6–70.9) | 5.0 (3.3–6.7) | 27.7 (24.2–31.2) |
| Middle | 64.2 (60.2–68.2) | 4.7 (2.8–6.6) | 31.1 (27.3–34.9) | 64.4 (60.6–68.2) | 4.7 (2.8–6.6) | 30.9 (27.3–34.5) |
| Richer | 72.5 (68.5–76.4) | 4.7 (3.0–6.5) | 22.8 (19.1–26.5) | 71.2 (67.3–75.2) | 4.8 (3.0–6.5) | 24.0 (20.3–27.7) |
| Richest | 71.7 (67.3–76.1) | 5.5 (3.1–8.1) | 22.8 (19.1–26.4) | 68.4 (63.7–73.0) | 5.6 (2.9–8.2) | 26.1 (22.1–30.1) |
| **B. Difference from the poorest quintile, percentage points (95% CI)** |  |  |  |  |  |  |
| Poorest (reference) | Ref. | Ref. | Ref. | Ref. | Ref. | Ref. |
| Poorer | 5.5* (0.6–10.5) | −0.5 (−2.8 to 1.8) | −5.1* (−10.1 to −0.1) | 3.8 (−0.9 to 8.4) | −0.4 (−2.6 to 1.8) | −3.3 (−7.9 to 1.2) |
| Middle | 3.3 (−2.2 to 8.9) | −0.8 (−3.3 to 1.8) | −2.6 (−8.0 to 2.8) | 0.9 (−4.3 to 6.2) | −0.7 (−3.3 to 1.9) | −0.2 (−5.2 to 4.9) |
| Richer | 11.6*** (6.0–17.2) | −0.7 (−3.1 to 1.7) | −10.8*** (−16.1 to −5.6) | 7.7** (2.3–13.2) | −0.7 (−3.1 to 1.8) | −7.1** (−12.2 to −2.0) |
| Richest | 10.8** (4.6–17.0) | 0.1 (−3.1 to 3.3) | −10.9*** (−16.1 to −5.6) | 4.9 (−1.6 to 11.3) | 0.1 (−3.3 to 3.6) | −5.0 (−10.6 to 0.6) |

*Note.* PNC = postnatal care. Predicted probabilities are survey-weighted marginal predictions from the multinomial models, expressed as percentages. Model 1 is adjusted for maternal age, birth order, marital status, residence and region; Model 4 additionally adjusts for maternal education, antenatal care visits and place of delivery. Panel B differences are relative to the poorest quintile; \* p < 0.05, \*\* p < 0.01, \*\*\* p < 0.001.

After adjustment for maternal education, ANC and place of delivery in Model 4, the wealth differences were attenuated. The predicted probability of timely PNC was 63.5% for the poorest group, compared with 71.2% for the richer group, a difference of 7.7 percentage points (95% CI 2.3–13.2). The corresponding difference for the richest group was 4.9 percentage points (95% CI −1.6 to 11.3) and was no longer statistically significant. For no PNC, the difference between the richer and poorest groups declined from −10.8 to −7.1 percentage points (95% CI −12.2 to −2.0), while the difference for the richest group declined to −5.0 percentage points (95% CI −10.6 to 0.6) and was no longer statistically significant.

Differences in delayed PNC remained small and statistically non-significant. Thus, adjustment reduced but did not eliminate the wealth gradient in PNC timing. The remaining difference was concentrated in the contrast between the poorest and richer groups, particularly for timely PNC and no PNC, rather than delayed PNC.

### Attenuation of wealth differences after adjustment

The average marginal effects were consistent with the multinomial results and further quantified the reduction in the wealth gradient after sequential adjustment (Table 5). For no PNC, women in the richer quintile had a 10.8 percentage-point lower predicted probability than women in the poorest quintile in Model 1 (95% CI −16.1 to −5.6), while the difference for the richest quintile was −10.9 percentage points (95% CI −16.1 to −5.6). These differences were reduced to −7.1 percentage points (95% CI −12.2 to −2.0) and −5.0 percentage points (95% CI −10.6 to 0.6), respectively, in Model 4. The corresponding attenuation was 34.5% for the richer group and 55.2% for the richest group. A similar pattern was observed when delayed and no PNC were combined as no timely PNC. In Model 1, the probability of no timely PNC was 11.5 percentage points lower among women in the richer quintile than among those in the poorest quintile (95% CI −17.1 to −5.8), and 10.8 percentage points lower among the richest group (95% CI −17.0 to −4.5). After full adjustment, these differences declined to −7.6 percentage points (95% CI −13.0 to −2.1) and −4.8 percentage points (95% CI −11.3 to 1.8), respectively. This corresponded to attenuation of 33.2% and 55.7%.

**Table 5.** Average marginal effects of household wealth quintile on the probability of no postnatal care and of no timely postnatal care, Models 1 to 4.

| **Household wealth quintile** | **Average marginal effect, percentage points (95% CI)** |  |  |  | **Attenuation, Model 1 to 4 (%)** |
|---|---|---|---|---|---|
|  | *Model 1* | *Model 2* | *Model 3* | *Model 4* |  |
| **A. No postnatal care** |  |  |  |  |  |
| Poorest (reference) | Ref. | Ref. | Ref. | Ref. | — |
| Poorer | −5.1 (−10.1 to −0.04)* | −4.5 (−9.4 to 0.4) | −4.2 (−8.9 to 0.6) | −3.3 (−7.9 to 1.3) | 32.6 |
| Middle | −2.6 (−7.9 to 2.8) | −1.6 (−6.8 to 3.7) | −1.4 (−6.7 to 3.8) | −0.1 (−5.2 to 4.9) | 92.4 |
| Richer | −10.8 (−16.1 to −5.6)*** | −9.1 (−14.4 to −3.8)** | −8.7 (−13.9 to −3.5)** | −7.1 (−12.2 to −2.0)** | 34.5 |
| Richest | −10.9 (−16.1 to −5.6)*** | −6.6 (−12.4 to −0.8)* | −6.4 (−12.1 to −0.6)* | −5.0 (−10.6 to 0.6) | 55.2 |
| **B. No timely postnatal care (delayed or none)** |  |  |  |  |  |
| Poorest (reference) | Ref. | Ref. | Ref. | Ref. | — |
| Poorer | −5.5 (−10.5 to −0.6)* | −4.9 (−9.7 to −0.04)* | −4.6 (−9.3 to 0.1) | −3.7 (−8.4 to 0.9) | 30.1 |
| Middle | −3.3 (−8.9 to 2.2) | −2.1 (−7.6 to 3.3) | −2.0 (−7.4 to 3.4) | −0.9 (−6.1 to 4.4) | 72.0 |
| Richer | −11.5 (−17.1 to −5.8)*** | −9.4 (−15.0 to −3.7)** | −9.0 (−14.5 to −3.4)** | −7.6 (−13.0 to −2.1)** | 33.2 |
| Richest | −10.8 (−17.0 to −4.5)** | −6.2 (−12.9 to 0.5) | −6.0 (−12.7 to 0.7) | −4.8 (−11.3 to 1.8) | 55.7 |

*Note.* Average marginal effects are expressed in percentage points relative to the poorest quintile. Models are as described in Tables 3 and 4. Attenuation is the percentage reduction in the quintile coefficient from Model 1 to Model 4, calculated from point estimates; confidence intervals are not shown. \* p < 0.05, \*\* p < 0.01, \*\*\* p < 0.001.

The reduction in the wealth differences occurred progressively as maternal education, ANC and place of delivery were added to the models. For the richer group, the difference in no PNC declined from −10.8 percentage points in Model 1 to −9.1 in Model 2, −8.7 in Model 3 and −7.1 in Model 4. The corresponding reductions for no timely PNC were from −11.5 to −9.4, −9.0 and −7.6 percentage points. The middle wealth group showed little evidence of a remaining difference after adjustment, while the poorer group had smaller differences that were no longer statistically significant in the fully adjusted model. The sequential models indicate that education, ANC and place of delivery accounted for part of the socioeconomic difference in PNC, but did not remove the difference between the poorest and richer groups. The attenuation was greater for the richest group, particularly for no PNC and no timely PNC, while the remaining gradient was concentrated mainly in the richer-versus-poorest comparison.

### Absolute and relative socioeconomic inequality in postnatal care

The SII and RII provided evidence of a wealth gradient across the full socioeconomic distribution (Table 6). In Model 1, the SII for no PNC was −15.2 percentage points (95% CI −21.5 to −8.9), indicating a lower probability of no PNC among women at higher positions in the wealth distribution. The corresponding RII was 0.58 (95% CI 0.47–0.73). For not timely PNC, the SII was −16.0 percentage points (95% CI −23.0 to −8.9) and the RII was 0.62 (95% CI 0.50–0.77). The inequality indices were attenuated as education, ANC and place of delivery were added to the models. For no PNC, the SII declined in magnitude from −15.2 percentage points in Model 1 to −11.0 in Model 2, −10.6 in Model 3 and −8.4 in Model 4.

**Table 6.**
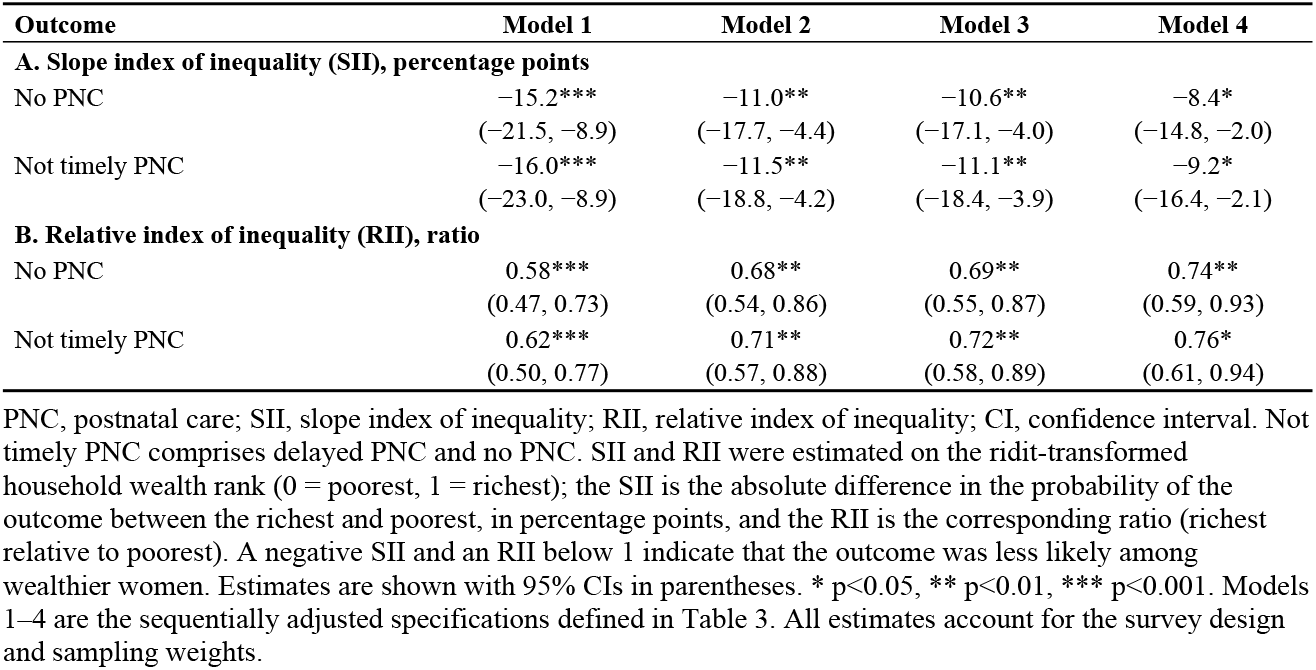
Wealth-related absolute (SII) and relative (RII) inequality in no postnatal care and in not timely postnatal care, Malawi Demographic and Health Survey 2024.

| **Outcome** | **Model 1** | **Model 2** | **Model 3** | **Model 4** |
|---|---|---|---|---|
| **A. Slope index of inequality (SII), percentage points** |  |  |  |  |
| No PNC | −15.2*** | −11.0** | −10.6** | −8.4* |
|  | (−21.5, −8.9) | (−17.7, −4.4) | (−17.1, −4.0) | (−14.8, −2.0) |
| Not timely PNC | −16.0*** | −11.5** | −11.1** | −9.2* |
|  | (−23.0, −8.9) | (−18.8, −4.2) | (−18.4, −3.9) | (−16.4, −2.1) |
| **B. Relative index of inequality (RII), ratio** |  |  |  |  |
| No PNC | 0.58*** | 0.68** | 0.69** | 0.74** |
|  | (0.47, 0.73) | (0.54, 0.86) | (0.55, 0.87) | (0.59, 0.93) |
| Not timely PNC | 0.62*** | 0.71** | 0.72** | 0.76* |
|  | (0.50, 0.77) | (0.57, 0.88) | (0.58, 0.89) | (0.61, 0.94) |

PNC, postnatal care; SII, slope index of inequality; RII, relative index of inequality; CI, confidence interval. Not timely PNC comprises delayed PNC and no PNC. SII and RII were estimated on the ridit-transformed household wealth rank (0 = poorest, 1 = richest); the SII is the absolute difference in the probability of the outcome between the richest and poorest, in percentage points, and the RII is the corresponding ratio (richest relative to poorest). A negative SII and an RII below 1 indicate that the outcome was less likely among wealthier women. Estimates are shown with 95% CIs in parentheses. \* p<0.05, \*\* p<0.01, \*\*\* p<0.001. Models 1–4 are the sequentially adjusted specifications defined in Table 3. All estimates account for the survey design and sampling weights.

The RII increased from 0.58 to 0.68, 0.69 and 0.74 across the same models. Nevertheless, both indices remained statistically significant in the fully adjusted model. A similar pattern was observed for not timely PNC, with the SII changing from −16.0 to −9.2 percentage points and the RII from 0.62 to 0.76 between Models 1 and 4. Thus, the inequality measures indicate that the wealth gradient was not limited to differences between individual wealth quintiles. Across the socioeconomic distribution, wealthier women had lower probabilities of no PNC and of not receiving PNC on time. Adjustment for maternal education, ANC and place of delivery reduced the magnitude of this inequality but did not eliminate it.

### Pathways linking household wealth to postnatal care

The pathway analysis examined whether antenatal care and delivery-facility type accounted for part of the association between household wealth and postnatal care outcomes (Table 7). Among all births, a one-quintile increase in household wealth was associated with a lower probability of no PNC by 1.40 percentage points (95% CI −2.68 to −0.13) after accounting for the specified pathways. The total effect was −1.91 percentage points (95% CI −3.25 to −0.57), with a total indirect effect of −0.51 percentage points (95% CI −0.83 to −0.19). This corresponds to 26.6% (95% CI 5.9–47.3) of the total association being mediated through the pathways included in the model. The significant indirect pathway was through home delivery. The indirect effect through home delivery was −0.42 percentage points (95% CI −0.72 to −0.11) for no PNC and −0.38 percentage points (95% CI −0.65 to −0.10) for not timely PNC. In contrast, the indirect effects through ANC alone were not statistically significant (−0.06 and −0.05 percentage points, respectively). The pathway operating through ANC followed by home delivery was also not statistically significant. For not timely PNC, the total indirect effect was −0.45 percentage points (95% CI −0.74 to −0.16), representing 23.5% (95% CI 3.6–43.3) of the total association.

**Table 7.**
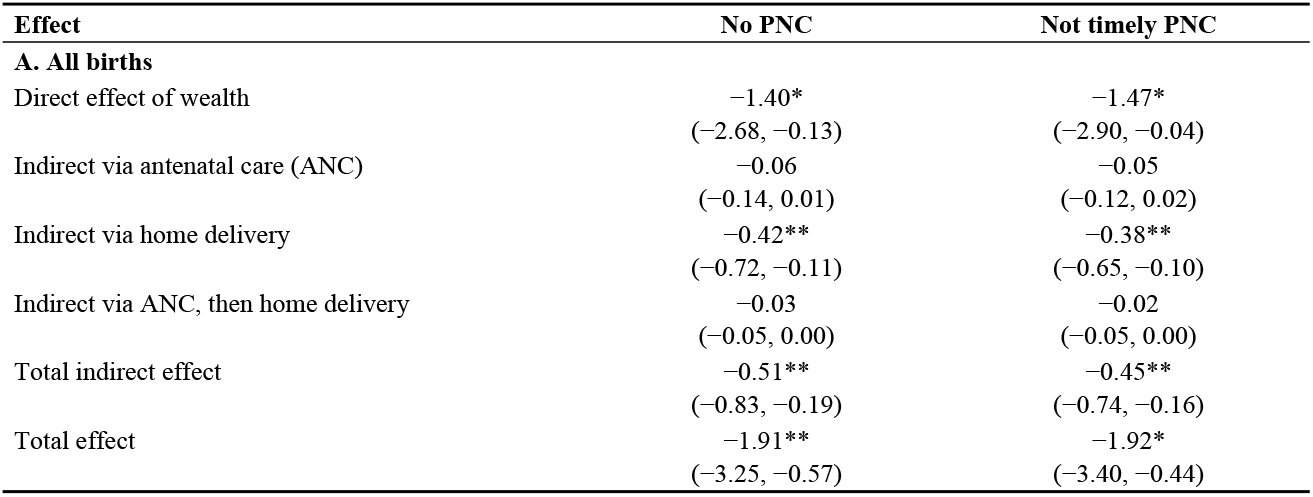
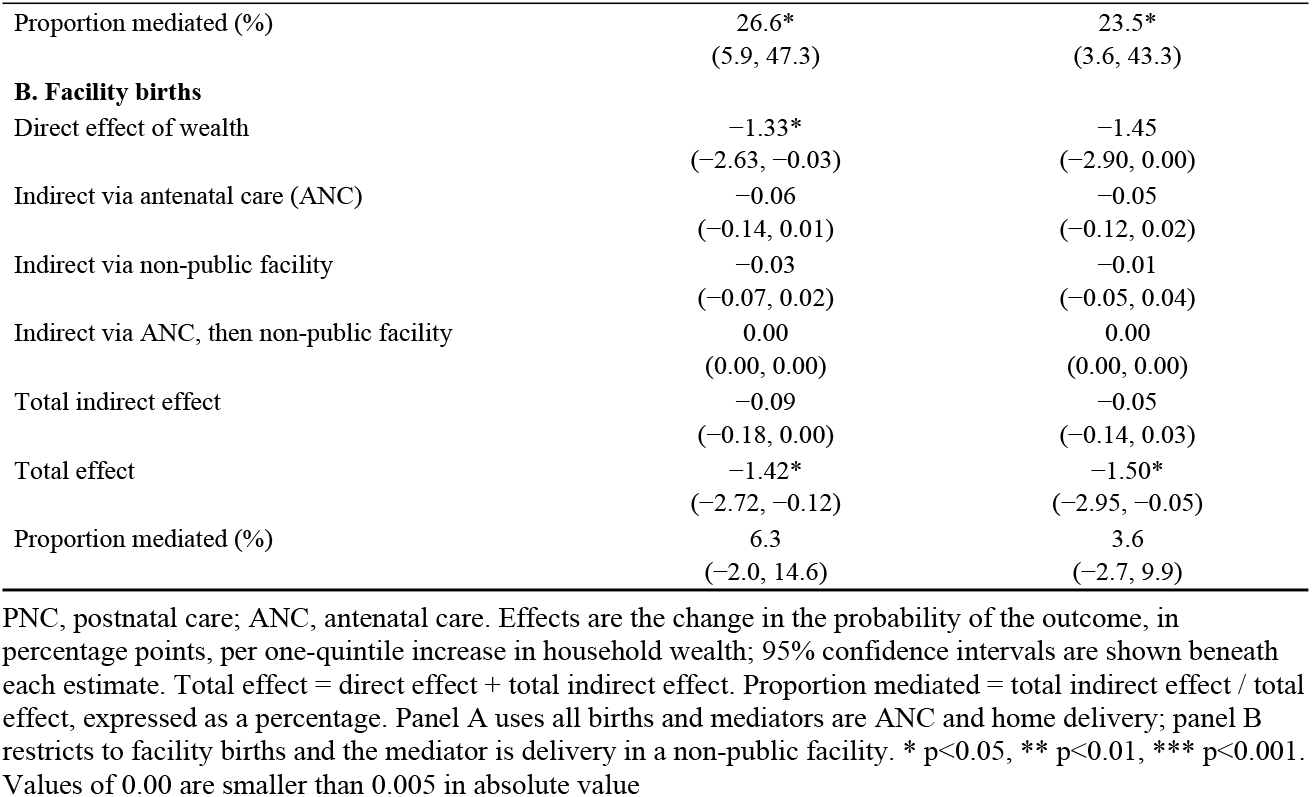
Direct and indirect (mediated) effects of household wealth quintile on postnatal care outcomes through antenatal care and delivery-facility type, Malawi Demographic and Health Survey 2024.

| Effect | No PNC | Not timely PNC |
|---|---|---|
| **A. All births** |  |  |
| Direct effect of wealth | −1.40* | −1.47* |
|  | (−2.68, −0.13) | (−2.90, −0.04) |
| Indirect via antenatal care (ANC) | −0.06 | −0.05 |
|  | (−0.14, 0.01) | (−0.12, 0.02) |
| Indirect via home delivery | −0.42** | −0.38** |
|  | (−0.72, −0.11) | (−0.65, −0.10) |
| Indirect via ANC, then home delivery | −0.03 | −0.02 |
|  | (−0.05, 0.00) | (−0.05, 0.00) |
| Total indirect effect | −0.51** | −0.45** |
|  | (−0.83, −0.19) | (−0.74, −0.16) |
| Total effect | −1.91** | −1.92* |
|  | (−3.25, −0.57) | (−3.40, −0.44) |
| Proportion mediated (%) | 26.6* | 23.5* |
|  | (5.9, 47.3) | (3.6, 43.3) |
| **B. Facility births** |  |  |
| Direct effect of wealth | −1.33* | −1.45 |
|  | (−2.63, −0.03) | (−2.90, 0.00) |
| Indirect via antenatal care (ANC) | −0.06 | −0.05 |
|  | (−0.14, 0.01) | (−0.12, 0.02) |
| Indirect via non-public facility | −0.03 | −0.01 |
|  | (−0.07, 0.02) | (−0.05, 0.04) |
| Indirect via ANC, then non-public facility | 0.00 | 0.00 |
|  | (0.00, 0.00) | (0.00, 0.00) |
| Total indirect effect | −0.09 | −0.05 |
|  | (−0.18, 0.00) | (−0.14, 0.03) |
| Total effect | −1.42* | −1.50* |
|  | (−2.72, −0.12) | (−2.95, −0.05) |
| Proportion mediated (%) | 6.3 | 3.6 |
|  | (−2.0, 14.6) | (−2.7, 9.9) |

PNC, postnatal care; ANC, antenatal care. Effects are the change in the probability of the outcome, in percentage points, per one-quintile increase in household wealth; 95% confidence intervals are shown beneath each estimate. Total effect = direct effect + total indirect effect. Proportion mediated = total indirect effect / total effect, expressed as a percentage. Panel A uses all births and mediators are ANC and home delivery; panel B restricts to facility births and the mediator is delivery in a non-public facility. \* p<0.05, \*\* p<0.01, \*\*\* p<0.001. Values of 0.00 are smaller than 0.005 in absolute value

The facility-birth analysis showed less evidence that delivery-facility type accounted for the wealth gradient. The total indirect effect was −0.09 percentage points (95% CI −0.18 to 0.00) for no PNC and −0.05 percentage points (95% CI −0.14 to 0.03) for not timely PNC. Neither the pathway through ANC nor that through delivery in a non-public facility was statistically significant. The corresponding proportions mediated were 6.3% (95% CI −2.0 to 14.6) for no PNC and 3.6% (95% CI −2.7 to 9.9) for not timely PNC. The pathway analysis indicates that the wealth gradient in PNC was partly accounted for by the pathway involving home delivery when all births were considered. This pathway was much weaker among facility births, where the estimated mediated proportions were small and their confidence intervals included zero. The results therefore point more clearly to delivery outside a health facility than to ANC or type of facility among facility births as a pathway through which household wealth was associated with PNC timing.

## Discussion

This study identified four main findings. First, although two-thirds of women received postnatal care within two days of childbirth, more than one-quarter received no postnatal care, indicating that substantial gaps remain despite the high level of facility delivery in Malawi. Second, postnatal care showed a clear socioeconomic gradient, with poorer women less likely to receive timely care and more likely to receive no care. Third, the wealth gradient was partly explained by maternal education, antenatal care and place of delivery, but remained after adjustment. Finally, home delivery accounted for a significant part of the wealth-related inequality in postnatal care, whereas antenatal care and type of facility among women who delivered in a facility explained little of the remaining inequality.

The proportion receiving timely postnatal care in Malawi was higher than that reported in several previous studies, although differences in definitions and timing criteria limit direct comparisons. For example (10), reported that only 16.5% of women in Ethiopia received postnatal care within two days, while (7) found that 52% of women in Bangladesh received postnatal care from a medically trained provider within two days in 2017. (6) also reported that only 41.4% of women received quality maternal postnatal care across 23 low- and middle-income countries. The relatively higher coverage observed in Malawi may partly reflect the high proportion of institutional births, which creates an opportunity for mothers and newborns to come into contact with health workers immediately after delivery. However, the finding that 28.3% received no postnatal care shows that contact with maternal health services before or during childbirth does not necessarily translate into postnatal follow-up.

Socioeconomic inequality was particularly evident in the difference between receiving timely care and receiving no care. Women in the poorest households had the highest prevalence of non-use, while women in richer households were more likely to receive timely care. Similar socioeconomic gradients have been reported in Bangladesh by (7), in Ethiopia by (25), and in India by (26). In the present study, however, wealth was not clearly associated with delayed postnatal care. This suggests that socioeconomic disadvantage may be more important in determining whether women receive postnatal care at all than in determining whether care is delayed once contact has been established. Financial constraints, transport costs, competing household responsibilities and the resources required to return to a health facility may make it particularly difficult for poorer women to initiate postnatal care after childbirth.

Maternal education and antenatal care were also strongly related to postnatal care. Women with higher education and those who attended ANC more frequently were more likely to receive timely postnatal care. (26) similarly reported positive associations between maternal education, household wealth and postnatal care in India, while (7) identified lack of ANC as an important characteristic among women who did not receive postnatal care in Bangladesh. Education may improve knowledge of maternal and newborn danger signs, understanding of the importance of postnatal checks and confidence in navigating health services. Antenatal care provides repeated opportunities to communicate this information and prepare women for care after delivery. However, the pathway analysis in this study showed that ANC alone did not significantly account for the wealth gradient. This suggests that having more ANC contacts does not necessarily ensure continuity into the postnatal period. The content of counselling, preparation for postnatal follow-up and the extent to which women are actively linked to services after delivery may be more important than the number of ANC contacts alone.

Place of delivery provided a clearer explanation of the socioeconomic difference.

Women who delivered at home were substantially less likely to receive timely postnatal care and were much more likely to receive no care than women who delivered in health facilities. Home delivery also represented the only statistically significant pathway through which wealth accounted for part of the difference in postnatal care, explaining approximately 27% of the total association with non-use. (16,27,28) similarly identified home delivery as an important barrier to timely postnatal care in Bangladesh. The explanation is plausible because facility delivery creates an immediate opportunity for maternal and newborn assessment, counselling and initiation of postnatal care before discharge, whereas women delivering at home must make a separate decision to seek care after childbirth. This additional step may be particularly difficult for women with fewer economic resources.

Importantly, however, the analysis restricted to facility births showed that socioeconomic inequality did not disappear. Among women who delivered in a health facility, the indirect pathway through facility type explained only a small and statistically non-significant proportion of the wealth gradient. This finding suggests that increasing institutional delivery, although important, may not be sufficient to eliminate socioeconomic differences in postnatal care. A similar pattern was reported by (26) in India, where socioeconomic differences in postnatal care persisted despite high levels of institutional delivery. Evidence from rural Malawi also provides a useful explanation. Kim et al. found that the availability of health facilities did not consistently translate into greater use of maternal and newborn postnatal care, suggesting that physical access alone does not guarantee utilisation. Differences in counselling, discharge practices, awareness of the need for follow-up, provider responsiveness and the ability to return to the facility may therefore become more important once women have overcome the initial barrier of reaching a facility.

The persistence of inequality after adjustment was also evident in the SII and RII estimates. Although adjustment for education, ANC and place of delivery reduced the magnitude of socioeconomic inequality, a significant gradient remained. This indicates that the observed differences were not confined to the poorest and richest households but extended across the socioeconomic distribution. Regional differences in postnatal care further suggest that household resources operate within a wider health-system and geographic context. (29) reported substantial geographic variation in the quality of newborn postnatal care in Ethiopia, while (26) found marked regional differences in postnatal care in India. In Malawi, regional differences may reflect variation in service availability, transport, health-system performance and the quality of postnatal care provided across facilities.

### Strengths and limitations

The use of the nationally representative 2024 Malawi Demographic and Health Survey allowed the analysis to examine socioeconomic inequalities in postnatal care at the population level. The study also combined multivariable regression with absolute and relative measures of inequality and pathway analysis, allowing the wealth gradient to be examined from different perspectives. The analysis further assessed whether the inequality persisted among women who delivered in a health facility, providing a useful comparison of the pathways operating before and after facility delivery. A key limitation was the cross-sectional nature of the data, which prevented us from establishing temporal or causal relationships between socioeconomic position, antenatal care, place of delivery and postnatal care.

Although the pathway analysis identified associations through ANC and delivery setting, these estimates should not be interpreted as causal mediation effects.

The use of secondary data also limited the analysis to variables collected in the MDHS. Important factors that may influence postnatal care, including the quality and content of counselling, discharge practices, provider behaviour, continuity of care and women’s experiences at health facilities, were not adequately captured. Information on ANC, place of delivery and timing of postnatal care was based on maternal report and may therefore be affected by recall or reporting error. Residual confounding is also possible despite adjustment for available sociodemographic and maternal characteristics. Finally, the decomposition and inequality analyses depend on the variables included in the models and should therefore be interpreted as statistical accounts of the observed wealth gradient rather than estimates of causal effects. Despite these limitations, the nationally representative sample and complementary inequality and pathway analyses provide useful evidence on the distribution of timely postnatal care in Malawi.

### Policy implications

The findings suggest that improving postnatal care in Malawi requires greater attention to the transition from pregnancy and childbirth to care after delivery. The strong association between home delivery and non-use of postnatal care, together with the significant pathway through home delivery, supports continued efforts to reduce home births and to strengthen mechanisms linking women who deliver outside health facilities to postnatal services. However, the persistence of socioeconomic inequality among women who delivered in a facility suggests that institutional delivery alone is unlikely to remove the gap. ANC and delivery care therefore provide important opportunities to prepare women for postnatal follow-up. Counselling should clearly communicate the timing and importance of postnatal care, while discharge procedures could include active referral or scheduling of follow-up where feasible. Community health workers may also have an important role in identifying mothers and newborns who do not receive timely care, particularly among poorer and rural households. Because education, ANC and place of delivery accounted for only part of the wealth gradient, efforts should also address barriers that arise after facility delivery, including the ability to return for follow-up and continuity between facility and community services. Monitoring postnatal care by socioeconomic position and region will be important to ensure that improvements in overall coverage do not conceal persistent inequalities.

## Conclusions

In Malawi, socioeconomic inequalities in postnatal care remain substantial, with poorer women more likely to receive no care and less likely to receive care within two days of childbirth. The wealth gradient was partly reduced after accounting for maternal education, ANC and place of delivery, but a significant inequality remained. Home delivery accounted for an important part of the wealth-related difference, whereas ANC and facility type explained little of the remaining inequality among women who delivered in health facilities. These findings suggest that increasing institutional delivery, although important, will not by itself eliminate socioeconomic inequalities in postnatal care. Strengthening the transition from ANC and childbirth to postnatal follow-up, together with targeted community and facility-based support for disadvantaged women, may help reduce the remaining gap.

Longitudinal studies are needed to clarify the temporal pathways underlying these inequalities and to assess which interventions are most effective in improving equitable postnatal care.

## Data Availability

The data used in this study are from the 2024 Malawi Demographic and Health Survey (MDHS). The dataset is publicly available through The DHS Program, subject to registration and approval of a data access request. Researchers can request access at https://dhsprogram.com/data/available-datasets.cfm.

## Supporting information

**S1 Table. Derivation of the analytic samples, 2024 Malawi Demographic and Health Survey**.

**S3 Table. Timing of the first postnatal check by background characteristics among facility births (n = 4**,**593)**.

**S4 Table. Distribution of the pathway variables by household wealth and place of delivery, all births (n = 4**,**726) except panel D**.

**S5 Table. Multinomial logistic regression of the timing of the first postnatal check, all births: full models 1–4**.

**S6 Table. Multinomial logistic regression of the timing of the first postnatal check, facility births: full models 1–4**.

**S7 Table. Distribution of the timing of the first postnatal check under alternative definitions, all births (n = 4**,**726)**.

**S10 Table. Sparse-cell checks: unweighted counts behind the pathway and facility-type results, all births (n = 4**,**726)**.

